# Dietary Supplement Counseling Practices Among Pharmacists in Lahore, Pakistan: Identifying Knowledge and Training Gaps

**DOI:** 10.64898/2026.07.22.26358717

**Authors:** Nida Bokharee, Nabeeha Naseer, Shayan Fatima, Amna Akbar, Rimshah Siddique, Shafaq Tajwar, Imran Waheed

## Abstract

**Background:** Dietary supplements (DS) are extensively used in Pakistan, frequently in conjunction with prescription medicines. Pharmacists are well positioned for patient counsesling due to their accessibility. However, their ability to provide evidence-based counseling remains uncertain due to their varying knowledge and training, especially in low- and middle-income countries (LMICs), including Pakistan. The study assessed pharmacists’ knowledge, attitudes, and practices (KAP) regarding DS counseling and identified gaps requiring intervention.

**Methods:** A cross-sectional study was conducted among 256 registered pharmacists in Lahore, Pakistan. Data were collected using a validated, self-administered questionnaire assessing knowledge, attitudes, and counseling practices concerning dietary supplements over a period of six months i.e., July to December 2025. Statistical analyses were performed using SPSS version 27. A *p-*value of < 0.05 was considered statistically significant.

**Results:** Mean age was 31.0 ± 6.2 years, with male predominance (56.3%), Doctor of Pharmacy degree (75.0%), community pharmacy practice (62.5%), and 1-5 years of experience (48.4%). Mean knowledge score was 6.18 ± 1.71, with most (76.6%) exhibiting moderate knowledge. Mean attitude score was 7.01 ± 1.94, with 91.4% demonstrating positive attitudes. Mean practice score was 7.91 ± 2.93, with 68% exhibiting good counseling practices. Knowledge scores were significantly higher among female pharmacists (6.96 ± 1.52 vs. 5.57 ± 1.61, p=0.001), urban residents (6.39 ± 1.61 vs. 4.37 ± 1.30, p<0.001), and clinical pharmacists (7.10 ± 1.68, p=0.001).

**Conclusion:** Pharmacists practicing in Lahore, Pakistan demonstrated positive attitudes but moderate knowledge and inconsistent counseling practices. Key gaps were identified in drug-supplement interaction knowledge and counseling practices. These findings underscore the need for organized education, continued training, and regulatory supervision to improve counseling practices of dietary supplements.

## INTRODUCTION

The global use of dietary supplements (DS) has increased, driven by growing interest in preventive health and self-care practices, often used in conjunction with conventional medicines. High DS consumption rates have been reported among various populations, notably multivitamins, omega-3 fatty acids, vitamin D, and probiotics for the prevention of chronic diseases and the maintenance of overall health [1]. Recently, increasing evidence supports consumer uptake of supplements across low- and middle-income countries (LMICs), where health information dissemination via digital media, increased urbanisation, and easy over-the-counter accessibility are reported causes [2, 3].

Accumulating evidence underscores potential hazards associated with excessive consumption, non-standardised formulations, and interactions with prescription medicines. These adverse outcomes contrast with the widespread perception that DS are inherently safe. Adverse outcomes, including compromised pharmacotherapy, hepatotoxicity, and bleeding disorders, have been reported, particularly with herbal preparations [4]. Moreover, pertinent safety concerns arise from drug–nutrient and herb–drug interactions, such as those between oral contraceptives or anticoagulants and St. John’s Wort, highlighting the need for professional supervision to ensure safe use of DS [5].

Pharmacists are often the first healthcare professionals to provide evidence-based advice when consulted by patients for DS counseling. While high-income countries are engaging pharmacists in expanded clinical roles, including independent prescribing and advanced patient care services, a significant portion of LMICs still lag in strengthening their pharmacy practice [6]. Existing studies indicate variability in pharmacists’ knowledge and practices regarding DS counseling [1, 7]. Moreover, these inconsistencies are evident from other LMICs including Malaysia, Iran, and Palestine, influenced by demographic and practice-related factors such as age and years of experience. A Malaysian study found that 73.5% of community pharmacists had average product knowledge, with gaps in dosing and referral practices. Similarly, a recent Iranian study reported that while 91.8% held positive attitudes, knowledge and practice scores were associated with age and experience. Another study conducted in Libya found above-average scores but low confidence (47.5%) in vitamin knowledge [5, 7-9]. These findings highlight persistent gaps in pharmacist preparedness across LMIC settings and underscore the need for organized education and professional development to provide effective supplemental counseling [6, 10].

In Pakistan, the use of supplements surged during the COVID-19 pandemic driven by products purporting to have immune-boosting properties. Despite this shift, studies reported inconsistent counseling practices and insufficient pharmacovigilance procedures among healthcare practitioners [11, 12]. Moreover, regulatory supervision of dietary supplements is limited, lack of standardised labels and with misleading advertisements posing potential health risks to consumers [11, 13]. Overall, there is a dearth of literature evaluating counseling practices of pharmacists on dietary supplements in Pakistan, limiting the ability to design targeted educational interventions and policy improvements. The Knowledge–Attitude–Practice (KAP) framework is the foundation of this investigation, which asserts that knowledge influences attitudes and consequently impacts practices.

Given the paucity of literature on pharmacists’ counseling practices and the increasing use of dietary supplements among the public in Pakistan, particularly within the Eastern Mediterranean region, the present study aimed to assess the knowledge, attitudes, and counseling practices, and demographic predictors, while addressing the critical gap in LMIC settings. The findings are expected to identify the gaps and areas for improvements in pharmacy education modules, continuing professional training, and regulatory initiatives aimed at strengthening the role of pharmacists in promoting rational and safe use of dietary supplements in LMICs.

## METHODS

### Ethical approval

The present study was carried out after obtaining ethical approval from the Institutional Review Board (IRB) of Akhtar Saeed College of Pharmacy (ASCP) in Lahore, Pakistan (Approval No.: ASCP-11/2025-Pharm.D-95). The current study was conducted in accordance with the principles of the Declaration of Helsinki. Written informed consent was obtained from all participants prior to the commencement of the study. Anonymity and confidentiality among the respondents were maintained throughout the study.

### Study design and duration

A descriptive, cross-sectional study was conducted among licensed pharmacists practicing in community, hospital, and clinical settings in Lahore, Pakistan. Data collection occurred over a six-months duration, i.e., July to December 2025. The present study was guided by Knowledge-Attitude-Practices (KAP) framework.

### Study population

The study population included licensed pharmacists involved in patient care and provision of pharmaceutical services.

**Inclusion criteria:** The present study included (1) registered pharmacists; (2) currently practicing in community, hospital, and clinical pharmacy settings; and (3) who provided voluntary informed consent to participate.**Exclusion criteria:** The target population who (1) failed to fulfil the inclusion criteria, (2) did not consent to participate, (3) were pharmacy technicians or assistants were not included in the study.

### Sample size and sampling technique

The sample size was estimated with Cochran’s formula. Due to the dearth of prior local data, the proportion (p) of pharmacists with potential knowledge or practice gaps was conservatively estimated at 26.5% [7]. The estimated sample size was 299, calculated at a 95% confidence level and a 5% margin of error. A two-stage sampling approach was used. First, a convenience sampling method was used to select community pharmacies, hospitals, and clinical pharmacy settings in Lahore were selected based on accessibility and willingness to participate. Second, a consecutive sampling method was used within these settings, all eligible pharmacists encountered during on-site visits were invited to participate until the target sample size was achieved. Of the 384 pharmacists approached, 256 completed the questionnaire (response rate: 66.6%), which closely approximates the calculated requirement. A detailed participant flow diagram is presented in *Fig. 1*.

**Fig. 1.**
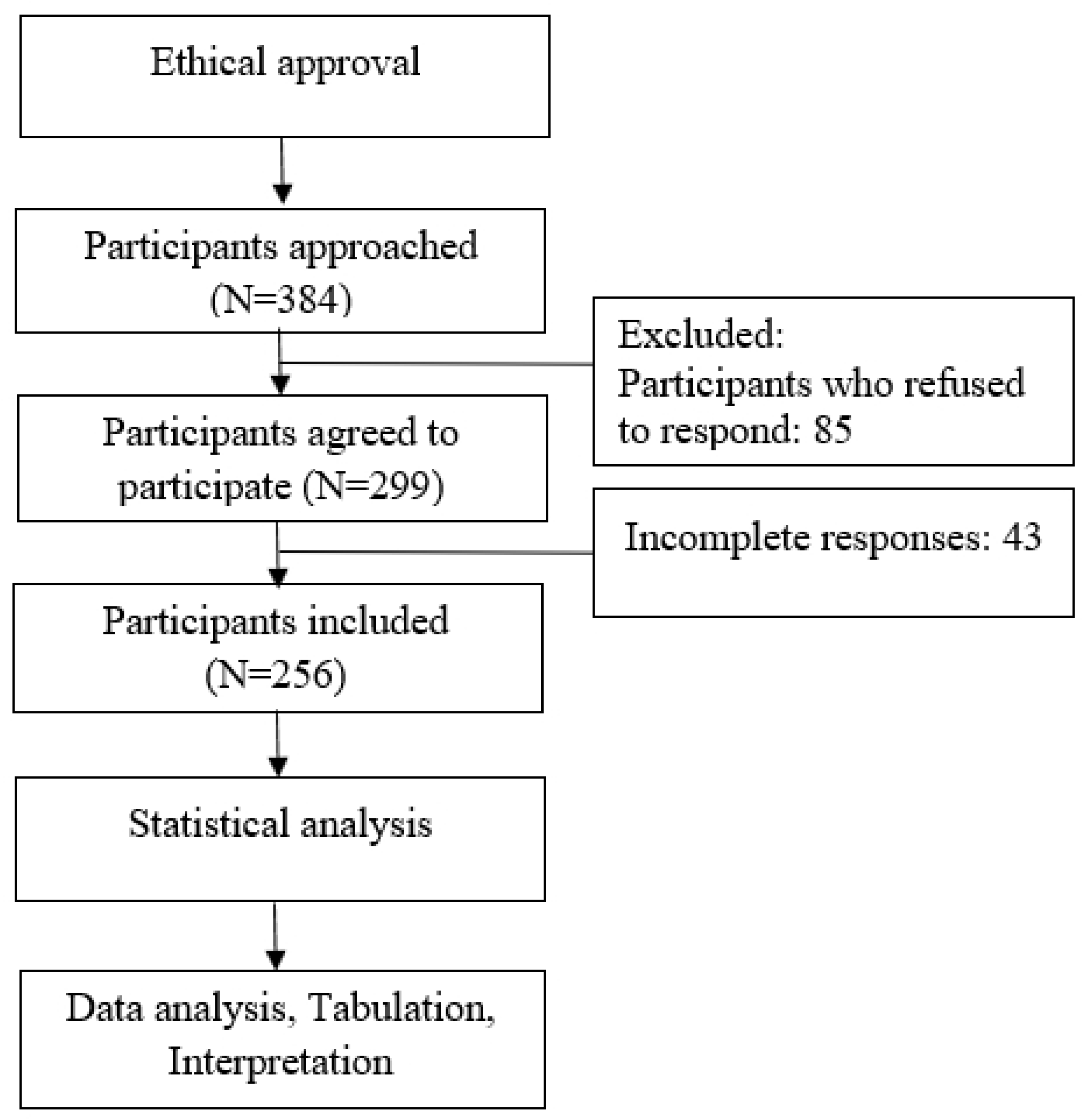
Study schematic diagram.

### Study tool

A structured, self-administered, paper-based questionnaire was designed following an extensive review of existing literature and tailored to the local context [7, 14-17]. Content validity was established via a panel of 6 expert pharmacists working in community, hospital, and clinical settings. The panel evaluated each item for relevance, clarity, and comprehensiveness, and their recommendations were incorporated. Face validity was tested through pilot testing (n = 35) with practicing pharmacists for clarity, and comprehension; these responses were not included in the final analyses. The internal consistency of the questionnaire was acceptable, with a Cronbach’s α value of 0.79. All the incomplete responses were also excluded from the final analyses.

The final questionnaire consisted of four sections, aligned with the KAP framework. Section I comprised questions regarding demographics such as age, gender, educational status, workplace setting, designation, and years of experience. Section II contained 11 close-ended questions evaluating the knowledge of the participants regarding dietary and nutritional supplements, including indications, benefits, side effects, and potential drug-nutrient interactions. Each correct answer was given 1 point, and an incorrect/uncertain answer received 0 points. A score of 9–11 on the knowledge section reflected good knowledge, followed by average knowledge with a score of 5–8, and poor knowledge with a score of 0–4. Section III comprised 9 items to assess the attitude of the study participants on a five-point Likert scale (strongly agree, agree, neutral, disagree, strongly disagree; 5 for strongly agree to 1 for strongly disagree) towards counseling on dietary supplements. Responses were categorized as positive and neutral/negative for reporting purposes. A total attitude score >5 was considered a positive attitude. Section IV comprises 11 statements about the practices of participants on a five-point Likert scale to assess self-reported counseling behaviors. Similarly, for reporting purposes, responses were categorized into positive and neutral/negative. A total practice score of >6 was considered good practice. The scoring system and cut-off points were based on a proportionate adaptation of previously published KAP studies in pharmacy practice [7].

### Data collection

Data were collected using paper-based, self-administered questionnaires. Researchers visited community pharmacies, hospitals, and clinical settings in Lahore and personally distributed questionnaires to eligible pharmacists who met the inclusion criteria. Written informed consent from the voluntary participants was obtained prior to data collection.

### Statistical analysis

Data were analyzed using Statistical Package for Social Sciences (SPSS) version 27. Continuous variables were summarized as mean ± SD, and categorical variables as frequencies and percentages, such as demographics and KAP scores. Normality of the data set was assessed using the Shapiro–Wilk test and visual inspection of Q–Q plots and histograms together. Independent t-test was used to compare the KAP scores between gender and residence. One-way ANOVA was used for comparisons across more than two groups such as age, education, experience, workplace. Linear regression was used to identify predictors of knowledge, attitude, and practice scores. A *p-* value < 0.05 considered statistically significant.

## RESULTS

### Sociodemographic characteristics of the study participants

A total of 256 pharmacists were included in the final analysis, with a 66.6% response rate. The mean age was 31.0 ± 6.2 years. The majority of the study participants were aged 25 to 39 years (n=172, 66.8%), more than half were male (n=144, 56.3%), and they resided in urban areas (n=229, 89.5%). A larger proportion held a Doctor of Pharmacy (Pharm. D) degree (n=192, 75%) and had 1–5 years of experience (n=124, 48.4%). The majority of pharmacists were employed in community pharmacies (n=160, 62.5%), followed by hospitals (n=83, 32.4%) and clinical pharmacy environments (n=13, 5.1%) (*Table 1*).

**Table 1.** Sociodemographic characteristics and Knowledge Attitude and Practice (KAP) scores of the study participant.

| Variable | n (%) | Knowledge Score<br>(Mean ± SD) | p-value<br>(Knowledge) | Attitude Score<br>(Mean ± SD) | p-value<br>(Attitude) | Practice Score<br>(Mean ± SD) | p-value<br>(Practice) |
| --- | --- | --- | --- | --- | --- | --- | --- |
| Mean age ± SD (years) | 31.0 ± 6.2 |  |  |  |  |  |  |
| Age (years) |  |  |  |  |  |  |  |
| <25 | 63 (24.6) | 5.81 ± 1.55 | 0.026* | 7.49 ± 1.84 | 0.045* | 7.12 ± 2.68 | 0.102* |
| 25–39 | 172 (66.8) | 6.14 ± 1.68 |  | 6.80 ± 1.92 |  | 7.87 ± 2.90 |  |
| 40–54 | 22 (8.6) | 6.91 ± 1.40 |  | 7.20 ± 2.01 |  | 8.30 ± 2.96 |  |
| Gender |  |  |  |  |  |  |  |
| Male | 144 (56.3) | 5.57 ± 1.61 | 0.001** | 6.98 ± 1.94 | 0.218** | 7.80 ± 2.88 | 0.307** |
| Female | 112 (43.8) | 6.96 ± 1.52 |  | 7.12 ± 1.89 |  | 8.02 ± 2.92 |  |
| Residence |  |  |  |  |  |  |  |
| Urban | 229 (89.5) | 6.39 ± 1.61 | 0.001** | 6.98 ± 1.93 | 0.218** | 7.92 ± 2.92 | 0.401** |
| Rural | 27 (10.5) | 4.37 ± 1.30 |  | 7.26 ± 1.88 |  | 7.75 ± 2.81 |  |
| Education |  |  |  |  |  |  |  |
| Bachelor of Pharmacy | 13 (5.1) | 6.31 ± 1.74 | 0.356* | 6.89 ± 1.91 | 0.072* | 7.85 ± 2.92 | 0.214* |
| Doctor of Pharmacy | 192 (75.0) | 6.09 ± 1.68 |  | 7.02 ± 1.94 |  | 7.91 ± 2.93 |  |
| Postgraduate (MS/M. Phil) | 51 (19.9) | 6.47 ± 1.72 |  | 6.80 ± 1.95 |  | 8.12 ± 2.96 |  |
| Experience (years) |  |  |  |  |  |  |  |
| ≤1 | 73 (28.5) | 5.80 ± 1.60 | 0.010* | 6.85 ± 1.88 | 0.109* | 7.87 ± 2.90 | 0.102* |
| 1–5 | 124 (48.4) | 6.20 ± 1.69 |  | 6.95 ± 1.94 |  | 7.91 ± 2.88 |  |
| 6–10 | 34 (13.3) | 6.45 ± 1.70 |  | 7.10 ± 1.97 |  | 8.00 ± 2.95 |  |
| 11–15 | 21 (8.2) | 6.50 ± 1.65 |  | 7.20 ± 2.01 |  | 8.10 ± 2.96 |  |
| >20 | 4 (1.6) | 7.00 ± 1.50 |  | 7.25 ± 1.85 |  | 8.30 ± 3.00 |  |
| Workplace |  |  |  |  |  |  |  |
| Community | 160 (62.5) | 5.80 ± 1.63 | 0.001* | 6.95 ± 1.92 | 0.314* | 7.85 ± 2.87 | 0.202* |
| Hospital | 83 (32.4) | 6.89 ± 1.60 |  | 7.05 ± 1.95 |  | 8.00 ± 2.93 |  |
| Clinical | 13 (5.1) | 7.10 ± 1.68 |  | 7.20 ± 1.94 |  | 8.15 ± 2.98 |  |
| Overall KAP score |  |  |  |  |  |  |  |
| Total knowledge score | 6.18 ± 1.71 |  | Total attitude score | 7.01 ± 1.94 | Total practice score | 7.91 ± 2.93 |  |
| Good knowledge | 20 (7.8) |  | Positive attitude | 234 (91.4) | Good practice | 174 (68) |  |
| Moderate knowledge | 196 (76.6) |  | Negative attitude | 22 (8.6) | Poor practice | 82 (32) |  |
| Poor Knowledge | 40 (15.6) |  |  |  |  |  |  |

**SD:** Standard deviation; **KAP:** Knowledge, attitude, and practice; **n:** Number of participants;
\* *p*-values calculated using one-way ANOVA for variables with >2 groups (age, education, experience, workplace);
\*\* *p*-values calculated using an independent t-test for variables with 2 groups (gender and residence). A *p*-value < 0.05 is statistically significant

### Knowledge of the study participants regarding dietary supplements

Overall, the mean knowledge score was 6.18 ± 1.71 out of 11, with the majority (n=196, 76.6%) indicating moderate knowledge (*Table 1*). Key misconceptions included the belief that supplements could replace a balanced diet (n=116, 45.3%) and limited understanding of clinically relevant drug–supplement interactions. Moreover, specific supplement knowledge varied, such as lutein being identified as beneficial for age-related macular degeneration by (n=160, 62.5%), glucosamine and chondroitin for joint health (n=177, 69.1%), and Ginkgo biloba’s cognitive benefits being acknowledged by 59%. Probiotic awareness was high (n=212, 82.8%), and 65.5% correctly identified the ideal timing of administration. There was limited knowledge regarding the significant drug-supplement interactions, such as the impact of St. John’s Wort and folates on oral contraceptives (n=86, 33.6% and n=79, 30.9%) and the impact of calcium and iron on antibiotic absorption (n=86, 33.6% and n=85, 33.2%). For therapeutic uses, ginseng, St. John’s Wort, and lavender were recognized for mild anxiety/stress (n=101, 39.5%; n=41, 16.0%; and n=80, 31.3%, respectively) and green tea extract for weight management (n=224, 87.5%) (*Table 2*).

**Table 2.** Knowledge of the study participants regarding dietary supplements.

| Item | Correct Response<br>n (%) |
| --- | --- |
| Primary purpose of nutritional supplements is to supplement a diet & provide additional nutrition | 116 (45.3) |
| Most commonly used to manage age-related macular degeneration (AMD); Lutein | 160 (62.5) |
| Common supplements used to maintain joint health and osteoarthritis; Glucosamine & chondroitin | 177 (69.1) |
| Best describes Gingko biloba's effect: Cognitive/Neuroprotective | 151 (59) |
| Are you familiar with probiotics and their potential benefits for gut health | 212 (82.8) |
| Optimal timing of probiotics | 168 (65.5) |
| Drug-supplement interactions reduce the efficacy of oral contraceptives; on St. John's Wort & folates | St. John's Wort: 86 (33.6) /<br>Folates: 79 (30.9) |
| Drug-supplement interactions reduce antibiotic absorption (especially tetracyclines). | Calcium: 86 (33.6) /<br>Iron: 85 (33.2) |
| Supplement most commonly used for weight loss; Green tea extract | 224 (87.5) |
| Herbal supplements to help manage mild anxiety and stress | St. John's Wort: 41 (16) /<br>Lavender: 80 (31.3)/<br>Ginseng: 101 (39.5) |
| Common side effects related to omega-3 supplements | Fishy aftertaste: 128 (50) /<br>Constipation: 79 (30.9) |

Frequencies and percentages of correct responses are summarized in the table.
For multiple correct responses possible, frequency and percentages reflect the proportion of participants identifying each correct response.

Multivariable linear regression model demonstrated that knowledge scores were significantly higher among female pharmacists (B = 1.09, *p < 0.001*), urban residents (B = –1.67, *p < 0.001*), older pharmacists (B = 0.47, *p = 0.024*), and those working in hospital/clinical settings (B = 0.35, *p = 0.044*). Hospital and clinical settings were combined in the regression model due to the small sample size of clinical pharmacists (n=13). Years of experience and educational qualifications were not significant predictors (*Table 5*).

### Attitude of the study participants toward dietary supplement counseling

Overall, the majority of the respondents (91.4%) reported a positive attitude toward DS counseling, as indicated by a mean score of 7.01±1.94 out of 9 (*Table 1*). Over half of the participants (62.5%) agreed that pharmacists play a vital role in patient counseling, while 68.4% emphasised that supplement counseling is equivalent to medicine counseling. Moreover, a considerable proportion (60.5%) acknowledged the need for acquiring additional training to enhance their skills (*Table 3*). Although demographic factors had a negligible influence on attitudes, age exhibited a slight significance, with pharmacists younger than 25 attaining the highest mean attitude score (*Table 1*). Regression analysis stated that demographics represented approximately 2.7% variance in the attitude scores (R² = 0.027, *p > 0.05*), indicating that attitudes were predominantly uninfluenced by individual characteristics (*Table 5*).

**Table 3.** Attitude of Pharmacists toward Dietary Supplements (N = 256)

| Statement | n (%) * |
| --- | --- |
| Pharmacists play a pivotal role in counseling | 160 (62.5) |
| Patients trust pharmacists' guidance more than others | 130 (50.8) |
| Comfortable discussing benefits/risks | 138 (53.9) |
| Importance of supplement counseling is equal to medication counseling | 175 (68.4) |
| Patients actively seek guidance on supplements | 118 (46.1) |
| Patients confused about supplement use | 100 (39.1) |
| Need for additional training on supplements | 155 (60.5) |

\* Positive responses of the participants who "Agree" or "Strongly Agree" on a 5-point Likert scale are included in the table.

### Counseling Practices of the Study Participants

The mean practice score was 7.91 ± 2.93; with 68% exhibiting good counseling practices, while 32% demonstrated inadequate performance (*Table 1*). These practices were assessing allergies (53.1%), acquiring patient histories (52.7%), patient referrals to physicians when necessary (66%), and educating patients on dosage of medications (68.4%). However, inadequacies were noted in counseling pertaining to the side effects and drug interactions (40.2%), provision of written materials (28.1%), staying updated on research (33.6%), and conducting follow-ups (43%) (*Table 4*).

**Table 4.** Practice of Pharmacists toward Dietary Supplements (N = 256)

| Statement | n (%) * |
| --- | --- |
| Always take patient history before recommending supplements | 135 (52.7) |
| Assess allergy status before recommendation | 136 (53.1) |
| Refer patients to physicians when necessary | 169 (66.0) |
| Educate patients on dosage/administration | 175 (68.4) |
| Allocate sufficient time for counseling | 144 (56.3) |
| Counsel on potential side effects & interactions | 103 (40.2) |
| Provide written educational material | 72 (28.1) |
| Keep updated with research | 86 (33.6) |
| Follow up with patients | 110 (43.0) |
| Address patient concerns & misconceptions | 128 (50.0) |
| Encourage patients to share supplement info with other HCPs | 135 (52.7) |

\* Positive responses of the participants who “Agree” or “Strongly Agree” on a 5-point Likert scale are included in the table

Regression analysis demonstrated that age, gender, residence, workplace, experience, and qualifications accounted only for 2.8% of the variance in practice scores (R² = 0.028, *p > 0.05*). Additionally, linear regression demonstrated that demographic characteristics accounted for a minor proportion of variance in practice scores (R² = 0.028, *p > 0.05*), with none were significant predictors (*Table 5*).

**Table 5.** Predictors of Knowledge, Attitude, and Practice of the study participants.

| Dependent Variable | Predictor | B | CI | p-value | R<sup>2</sup> (Overall) |
| --- | --- | --- | --- | --- | --- |
| Knowledge | Gender (Female) | 1.09 | 0.65 - 1.53 | <0.001* | 0.284 |
|  | Residence (Urban) | -1.67 | -2.24 – (-1.10) | <0.001* |  |
|  | Workplace (Hospital/Clinical) | 0.35 | 0.01 - 0.69 | 0.044* |  |
|  | Age | 0.47 | 0.06 - 0.88 | 0.024* |  |
|  | Qualification | NS | - | 0.613 |  |
|  | Experience | NS | - | 0.801 |  |
| Attitude | Age (<25 highest) | 0.49 | -0.05 - 1.03 | 0.072 | 0.027 |
|  | Gender, Residence, Workplace, Experience, Qualification | NS |  | >0.05 |  |
| Practice | Age (positive trend) | 0.68 | -0.05 - 1.03 | 0.102 | 0.028 |
|  | Other demographics | NS |  | >0.05 |  |

**B:** Unstandardised regression coefficient; **CI:** 95% confidence interval for B; **NS:** Not significant; **R<sup>2</sup>** = Proportion of variance in the dependent variable explained by the model. \* The *p-value* for the regression model is statistically significant at < 0.05

## DISCUSSION

The increasing use of dietary supplements in Lahore, Pakistan, highlights the importance of ensuring that pharmacists are well-equipped to provide evidence-based counseling. The current study assessed the knowledge, attitude, and counseling practices of pharmacists regarding DS and examined associate predictors. Pharmacists demonstrated moderate knowledge and positive attitudes, but inconsistent counseling practices. These findings corroborate recent international as well as LMIC studies, indicating persistent knowledge gaps despite positive attitudes [1, 7, 8]. However, the presence of such gaps is notable given the rising consumption of dietary supplements in LMICs, including Pakistan, and the potential for clinically significant adverse effects and drug-nutrient interactions [4, 5]. The discrepancy between positive attitudes and inconsistent counselling practices suggests that favorable perceptions alone may not be sufficient to ensure consistent evidence-based counselling in routine pharmacy practice.

The moderate knowledge scores observed in our study are consistent with previous findings from Pakistan, reporting mixed levels of awareness among community pharmacists regarding supplements [11]. Similarly, another Pakistani study reported that pharmacists had a knowledge score of only 21.99% regarding complementary and alternative medicine, highlighting persistent knowledge gaps [18]. These findings collectively suggest that pharmacist knowledge regarding dietary supplements and related products remains limited in Pakistan, underscoring the need for targeted educational interventions.

Higher knowledge scores were observed among the female pharmacists, practitioners in urban regions, and those practicing in hospital or clinical settings consistent with the evidence that workplace and resource availability influence performance and practices [7, 8]. Additionally, the association between age and knowledge also corroborates the findings of previous studies [1, 8]. However, years of professional experience was not a significant predictor in the current study.

Although pharmacists reported positive attitude to participate in DS counseling; previous studies have identified limited training, lack of ongoing professional education, and low confidence as barriers to effective counseling [1, 4, 7]. The low proportion of variance explained by the demographic characteristics (R² = 2.7–2.8%) is suggestive that factors beyond individual’s demographics such as institutional support, workload, and ongoing education, may influence attitude and counseling practices [10-12]. These observations provide a possible explanation for the knowledge and practice gaps observed in the present study.

The present study identified moderate knowledge and specific misconceptions such as perceiving dietary supplements as substitutes for a balanced diet and lacking understanding of drug–nutrient interactions. This highlights the need for targeted educational initiatives. These findings are in line with recent studies showing that customised training such as a nutrition education course may contribute significantly to improve pharmacists’ nutrition knowledge and counseling practices [19].

From a pharmacy practice perspective, the findings of the present study highlight the gaps in pharmacists’ positive attitude to provide comprehensive counseling on DS. To strengthen pharmacists’ competencies in DS counseling, consideration of integrating dedicated DS and associated patient counseling into undergraduates’ pharmacy curricula. Moreover, this should be complemented by the continuing professional development programmes and post-training competency evaluations by professional bodies and employers. Integrating such training modules in the routine pharmacy services may contribute to improved therapeutic outcomes and overall patient safety. Regulatory supervision also remains essential in Pakistan, where inconsistent product standardisation and misleading advertisement persist [13]. Regulatory bodies, such as the Drug Regulatory Authority of Pakistan (DRAP), should strengthen oversight of DS marketing and labelling to ensure safe use.

However, several limitations should be considered. Firstly, a cross-sectional study limits causal inferences. Secondly, self-administered questionnaire may introduce response and recall bias. Thirdly, the convenience sampling technique and moderate response rate (66.6%) may limit generalisability. Fourthly, the current study was conducted in a single metropolitan city; therefore, the findings should be interpreted within this context and may not be generalizable to pharmacists practising in other regions of Pakistan. Lastly, the self-reported nature of the practice section may overestimate actual counseling behaviors due to social desirability bias. Furthermore, future studies may benefit from additional validation of the study tool in larger and more diverse populations. Despite these limitations, the study has several strengths. To our knowledge, this is one of the first studies in Pakistan to simultaneously evaluate pharmacists’ knowledge, attitudes, and counseling practices regarding DS across pharmacists working in different setups including community, hospital and clinical settings. Furthermore, the findings focus on the practicing pharmacists in a metropolitan city, highlighting their vital role in providing extended pharmacy services. However, future studies should utilise longitudinal and interventional study designs involving pharmacists from multiple regions of Pakistan, to assess the impact of pharmacist-led counseling on patient outcomes and inform strategies for promoting rational supplement use.

## CONCLUSIONS

Pharmacists in Lahore demonstrated moderate knowledge and positive attitudes toward dietary supplement counseling. However, knowledge and practices gaps were identified, highlighting areas requiring targeted educational interventions. While demographic factors were significant predictors of knowledge, they were not associated with attitudes or practices. These findings underscore the need for focused educational initiatives and continuing professional training and appropriate regulatory supervision to strengthen evidence-based dietary supplement counseling by pharmacists.

## Data Availability

The anonymized dataset and supporting materials are available from the corresponding author upon reasonable request.

## Acknowledgments

The authors would like to thank all the participants for taking out time and completing the survey for this study.

## Author contributions (CRediT)

Conceptualisation: N.B., N.N., S.F.

Project administration: N.B.; Data curation: N.N., S.F., A.A., R.S., S.T.

Formal analysis: N.B., N.N., I.W.

Methodology: N.B., I.W.

Investigation: N.N., S.F., A.A., R.S., S.T.

Supervision: N.B.

Writing original draft: N.N., S.F., A.A., R.S., S.T.

Writing review & editing: N.B., N.N., S.F., A.A., R.S., S.T.

## REFERENCES

1. Slavcheva K SR, Neycheva N, Kafalova D. Community Pharmacists’ Knowledge, Attitudes, and Readiness to Provide Counseling on Food Supplements—A Scoping Review. Nutrients. 2025;17(23):3754.

2. Lwakatare M MJ. Dietary supplement use and associated factors among adults working in urban settings in Tanzania: A cross-sectional study. Health Serv Insights. 2023;16:11786329231170752.

3. Tareq MA EU, Banna MHA, Rezyona H, Seidu AA, Abid MT, et al. Prevalence and factors associated with dietary supplement use among Bangladeshi public university students: A cross-sectional study. PLoS One. 2022;17(10):e0276343.

4. Stayduhar JM CJ, Schreiber JB, Witt-Enderby PA. Pharmacist and student knowledge and perceptions of herbal supplements and natural products. Pharmacy. 2023;11(3):96.

5. Balaban U YBT, Kelleci Cakir B. Assessment of food-drug interaction knowledge among Turkish healthcare professionals. BMC Med Educ. 2025;25(1):934.

6. ZUD B. Ten recommendations to improve pharmacy practice in low and middle-income countries (LMICs). J Pharm Policy Pract. 2021;14(1):6.

7. Xin RKW YT, Qin WZ, Kaiyee L, Mohammed AH, Blebil A, et al. Community pharmacists’ knowledge, attitudes, and practices regarding counselling on vitamins and dietary supplements in Malaysia: A study on complementary medicines. Explor Res Clin Soc Pharm. 2024;13:100410.

8. Bidoki MZ MM, Afzal G, Aghabagheri M, Meybodi MN. Knowledge, Attitude, and Practice Analysis of Dietary Supplements Among Pharmacists: A Promising Outlook from Yazd, Iran. J Res Pharm Pract. 2025;14(1):27–34.

9. Altamimi M HM, Badrasawi M, Allahham S. Knowledge, attitudes and practices related to dietary supplements among a group of Palestinian pharmacists. Sultan Qaboos Univ Med J. 2021;21(4):613.

10. Ng JY TU, Dhaliwal S. Barriers, knowledge, and training related to pharmacists’ counselling on dietary and herbal supplements: a systematic review of qualitative studies. BMC Health Serv Res. 2021;21(1):499.

11. Akhtar MM AA, Aqeel MT, Ullah M, Rana SM, Sufiyan B. Assessment of Knowledge, Attitudes, and Perceptions of Community Pharmacists Regarding Vitamin D and Calcium Products: A Qualitative Study in Islamabad and Rawalpindi, Pakistan. Curr Pharm Res. 2025;3(1):108–27.

12. Hashmi F HM, Saleem F, Saeed H, Islam M, Malik UR, et al. Perspectives of community pharmacists in Pakistan about practice change and implementation of extended pharmacy services: a mixed method study. Int J Clin Pharm. 2021;43(4):1090–100.

13. Mustafa ZU SM, Asif N, Rao AZ, Khan QUA, Nawaz AS, et al. Knowledge, attitude, practices, and barriers of pharmacovigilance among healthcare workers: a cross-sectional survey from Lahore, Pakistan. Bull Fac Pharm Cairo Univ. 2021;59(1):33–43.

14. Mehralian G YN, Hashemian F, Maleksabet H. Knowledge, attitude and practice of pharmacists regarding dietary supplements: a community pharmacy-based survey in Tehran. Iran J Pharm Res. 2014;13(4):1457.

15. Medhat M SN, Ashoush N. Knowledge, attitude and practice of community pharmacists towards nutrition counseling. Int J Clin Pharm. 2020;42(6):1456–68.

16. Ashiq K QM, Bajwa MA, Ashiq S, Abid F, Yasir S. Assessment of pharmacists towards the use of herbal medicines in Lahore, Pakistan. Rawal Med J. 2022;47(3):684.

17. Khan A AM, Aldarmahi A, Zaidi SF, Subahi AM, Al Shaikh A, et al. Awareness, self-use, perceptions, beliefs, and attitudes toward complementary and alternative medicines (CAM) among health professional students in King Saud bin Abdulaziz University for Health Sciences Jeddah, Saudi Arabia. Evid Based Complement Alternat Med. 2020;2020(1):7872819.

18. Tahir AH TM, Shahnaz G, Saqlain M, Ayub S, Ahmed A. Knowledge, attitude, and perceptions of healthcare professionals towards complementary and alternative medicine: a cross-sectional survey from twin cities of Pakistan. BMC Complement Med Ther. 2023;23(1):432.

19. Mirkazemi C WM, Berbecaru M, Stubbings T, Murray S, Veal F, et al. Practising pharmacists want more nutrition education. Curr Pharm Teach Learn. 2022;14(11):1420–30.

